# Ultra-fast and onsite interrogation of Severe Acute Respiratory Syndrome Coronavirus 2 (SARS-CoV-2) in environmental specimens via surface enhanced Raman scattering (SERS)

**DOI:** 10.1101/2020.05.02.20086876

**Authors:** Dayi Zhang, Xiaoling Zhang, Rui Ma, Songqiang Deng, Xinquan Wang, Xian Zhang, Xia Huang, Yi Liu, Guanghe Li, Jiuhui Qu, Yu Zhu, Junyi Li

**Author notes:** **Corresponding author:** Dr Dayi Zhang, School of Environment, Tsinghua University, Beijing 100084, P.R. China, Dr Junyi Li, Suzhou Yiqing Environmental Science and Technology LTD., Suzhou 215163, P.R. China.

## Abstract

The outbreak of coronavirus infectious disease-2019 (COVID-19) pneumonia challenges the rapid interrogation of the severe acute respiratory syndrome coronavirus 2 (SARS-CoV-2) in human and environmental specimens. In this study, we developed an assay using surface enhanced Raman scattering (SERS) coupled with multivariate analysis to diagnose SARS-CoV-2 in an ultra-fast manner without any pretreatment (e.g., RNA extraction). Using silver-nanorod SERS array functionalized with cellular receptor angiotensin-converting enzyme 2 (ACE2), we obtained strong SERS signals of ACE2 at 1032, 1051, 1089, 1189, 1447 and 1527 cm^−1^. The recognition and binding of receptor binding domain (RBD) of SARS-CoV-2 spike protein on SERS assay significantly quenched the spectral intensities of most peaks and exhibited a shift from 1189 to 1182 cm^−1^. On-site tests on 17 water samples with a portable Raman spectrometer proved its accuracy and easy-operation for spot diagnosis of SARS-CoV-2 to evaluate disinfection performance, explore viral survival in environmental media, assess viral decay in wastewater treatment plant and track SARS-CoV-2 in pipe network. Our findings raise a state-of-the-art spectroscopic tool to screen and interrogate viruses with RBD for human cell entry, proving its feasibility and potential as an ultra-fast diagnostic tool for public health.

## 1. Introduction

The outbreak of coronavirus infectious disease-2019 (COVID-19) pneumonia since 2019 is caused by the severe acute respiratory syndrome coronavirus 2 (SARS-CoV-2) ^1, 2, 3^ and it has rapidly spread throughout 202 countries around the world. Till 19^th^ April 2020, there have been over 2 million confirmed cases and 220,000 deaths globally, and the number is still increasing rapidly. As there is clear evidence of human-to-human transmission of SARS-CoV-2 ^2, 4, 5, 6^, e.g., direct contact, respiratory droplets ^3, 7, 8^ and stools ^9, 10, 11, 12, 13, 14^, how to interrogate SARS-CoV-2 in human and environmental specimens draws more attentions for effectively confirming COVID-19 cases and identifying transmission routes. It brings urgent requirement of developing diagnostic tools that can rapidly and specifically recognize SARS-CoV-2 in tracking patients.

Many approaches can detect SARS-CoV-2 with high specificity, e.g., real-time reverse transcription quantitative polymerase chain reaction (RT-qPCR) and serological enzyme-linked immunosorbent assays (ELISA). RT-qPCR targeted viral specific RNA fragment with specific primers for the open reading frame 1ab (CDDC-ORF), nucleocapsid protein (CDDC-N), envelope protein, membrane protein, or RNA-dependent RNA polymerase (RdRp)^15, 16, 17, 18^ RNA extraction from swab samples is necessary for RT-qPCR and requires time-consuming pretreatment normally taking more than 4 hours^19, 20^, bringing a barrier for rapid diagnosis of SARS-CoV-2. Alternatively, ELISA is a commonly used an enzyme immunoassay to detect a receptor using antibodies directed against the antigen ^21^, targeting the immunological markers, IgM and IgG antibodies, which are reported to increase in the blood of most patients more than a week after infection^22^. However, this method is still time-consuming and not feasible for diagnosing SARS-CoV-2 in environmental media which do not have immunological markers^23, 24, 25^. It is of great urgency to develop a rapid, reproducible, cheap and sensitive assay detecting SARS-CoV-2, especially applicable for different specimens.

Raman spectroscopy is a vibrational spectroscopy of ability to detect chemical bonds *via* photon scattering, but the generated signals are extremely weak comparing to the incident beam. Thus, surface enhanced Raman scattering (SERS) is introduced to overcome such inherent limitation and interrogate trace materials by exploiting the enormous electromagnetic field enhancement resulted from the excitation of localized surface plasmon resonances at nanostructured metallic surfaces, mostly gold or silver^26, 27^. It has been widely applied for biological analysis, *e.g*., living cell classification^28^, cancer detection^29^, biological imaging^30^ and virus detection^31^. For SARS-CoV-2, the spike glycoprotein consists of S1 and S2 subunits, and S1 subunit contains a receptor binding domain (RBD) directly recognizing the human receptor angiotensin converting enzyme 2 (ACE2) for cell entry ^32, 33^ Such recognition and binding might alter the structure of ACE2 and lead to changes in Raman spectra. Additionally, the binding specificity allows ACE2 as an anchor to capture SARS-CoV-2 from human or environmental specimens for interrogation.

In this study, we proposed a ‘capture-quenching’ strategy to rapidly detect SARS-CoV-2 and developed a SERS assay introducing ACE2 functionalized on silver-nanorod SERS substrates to capture and interrogate SARS-CoV-2 spike protein (Figure 1). The induced SERS signal quenching was documented by either red-shift or whole spectral alterations in multivariate analysis as biomarkers for the presence of SARS-CoV-2 in real environmental specimens.

**Figure 1.**
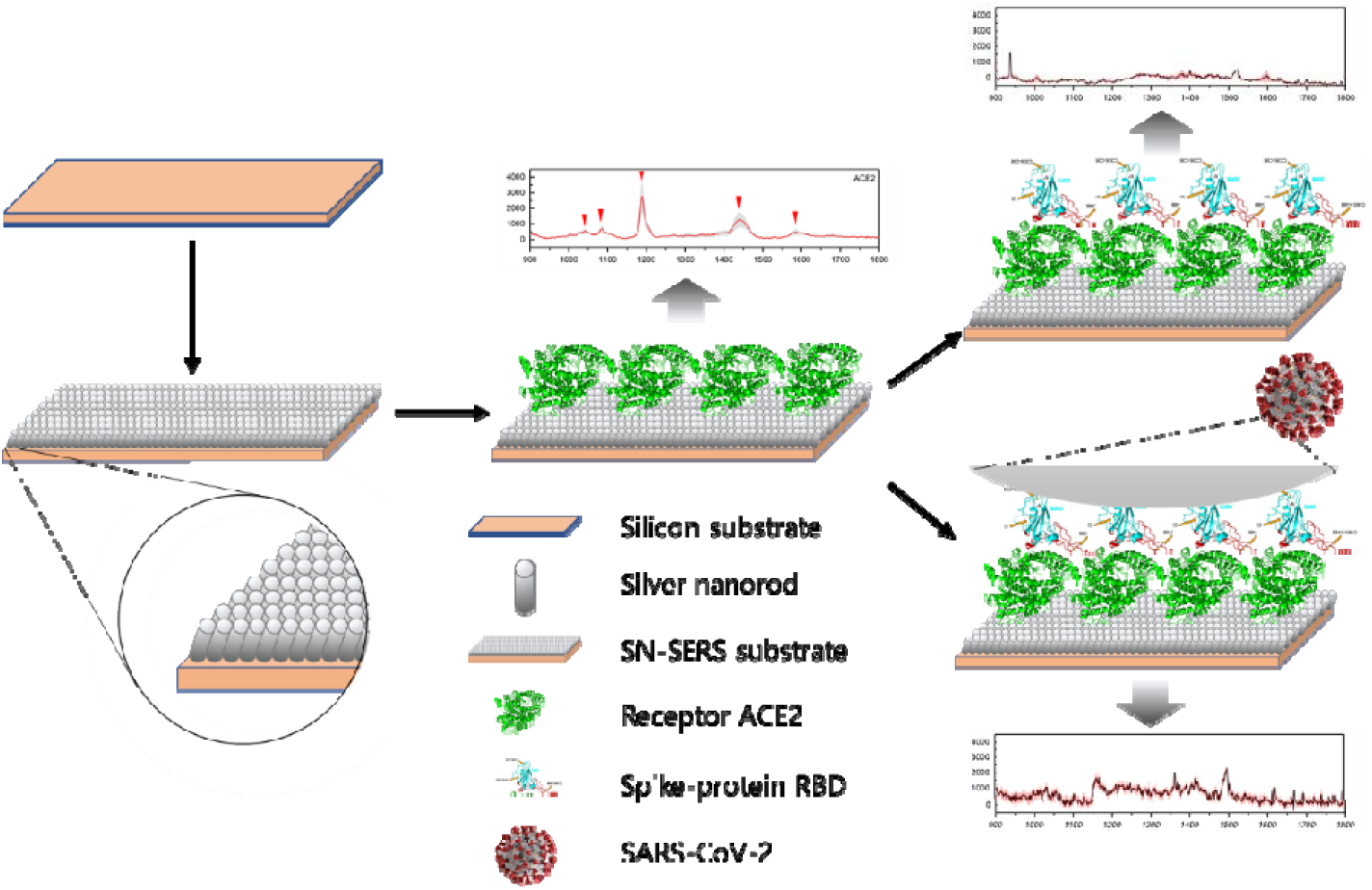
State-of-the-art diagram of surface enhanced Raman scattering (SERS) for interrogating Severe Acute Respiratory Syndrome Coronavirus 2 (SARS-CoV-2). Human cellular receptor Angiotensin-Converting Enzyme 2 (ACE2) is functionalized on silver-nanorod SERS (SN-SERS) substrate, designated as ACE2@SN-SERS array, generated strong SERS signals (1032, 1051, 1089, 1189, 1447 and 1527 cm^−1^). The recognition and binding of receptor binding domain (RBD) of SARS-CoV-2 spike protein on ACE2@SN-SERS assay significantly quenches the spectral intensities of most peaks and exhibits a red-shift from 1189 to 1182 cm^−1^.

## 2. Materials and methods

### 2.1 Water samples and biological analysis

Seventeen water samples were collected from hospitals and pipe network in Wuhan (China) from 24^th^ March to 10^th^ April, 2020 (Table 1). Around 2.0 L of water was directly collected in a plexiglass sampler, placed in 4°C ice-boxes and immediately transferred into laboratory for RNA extraction following our reported protocol ^34^ Briefly, after centrifugation at 3,000 rpm to remove suspended solids, the supernatant was subsequently supplemented with NaCl (0.3 mol/L) and PEG-6000 (10%), settled overnight at 4°C, and centrifuged at 10,000 g for 30 minutes. Viral RNA in pellets was extracted using the EZ1 virus Mini kit (Qiagen, Germany) according to the manufacturer’s instructions. SARS-CoV-2 RNA was quantified by RT-qPCR using AgPath-ID™ One-Step RT-PCR Kit (Life Technologies, USA) on a LightCycler 480 Real-time PCR platform (Roche, USA) in duplicates. Two target genes simultaneously amplified were open reading frame lab (CCDC-ORF1, forwards primer: 5‘-CCCT GT GGGTTTTACACTTAA-3’; reverse primer: 5‘-ACGATTGTGCATCAGCTGA-3’; fluorescence probe: 5‘-FAM-CCGTCTGCGGTATGTGGAAAGGTTATGG-BHQ1-3’) and nucleocapsid protein (CCDC-N, forwards primer: 5‘-GGGGAACTTCTCCTGCTAGAAT-3’; reverse primer: 5‘-CAGACATTTTGCTCTCAAGCTG-3’; fluorescence probe: 5‘-FAM-TTGCTGCTGCTTGACAGATT-TAMRA-3’). RT-qPCR amplification for CCDC-ORF1 and CCDC-N was performed in 25 μL reaction mixtures containing 12.5 μL of 2×RT-PCR Buffer, 1 μL of 25×RT-PCR Enzyme Mix, 4 μL mixtures of forward primer (400 nM), reverse primer (400 nM) and probe (120 nM), and 5 μL of template RNA. Reverse transcription was conducted at 45°C for 10 min (1 cycle), followed by initial denaturation at 90°C for 10 min (1 cycle) and 40 thermal cycles of 60°C for 45 second and 90°C for 15 seconds. Quantitative fluorescent signal for each sample was normalized by ROX™ passive reference dye provided in 2×RT-PCR buffer. For each RT-qPCR run, both positive and negative controls were included. For quality control, a reagent blank and extraction blank were included for RNA extraction procedure and no contamination was observed.

**Table 1.**
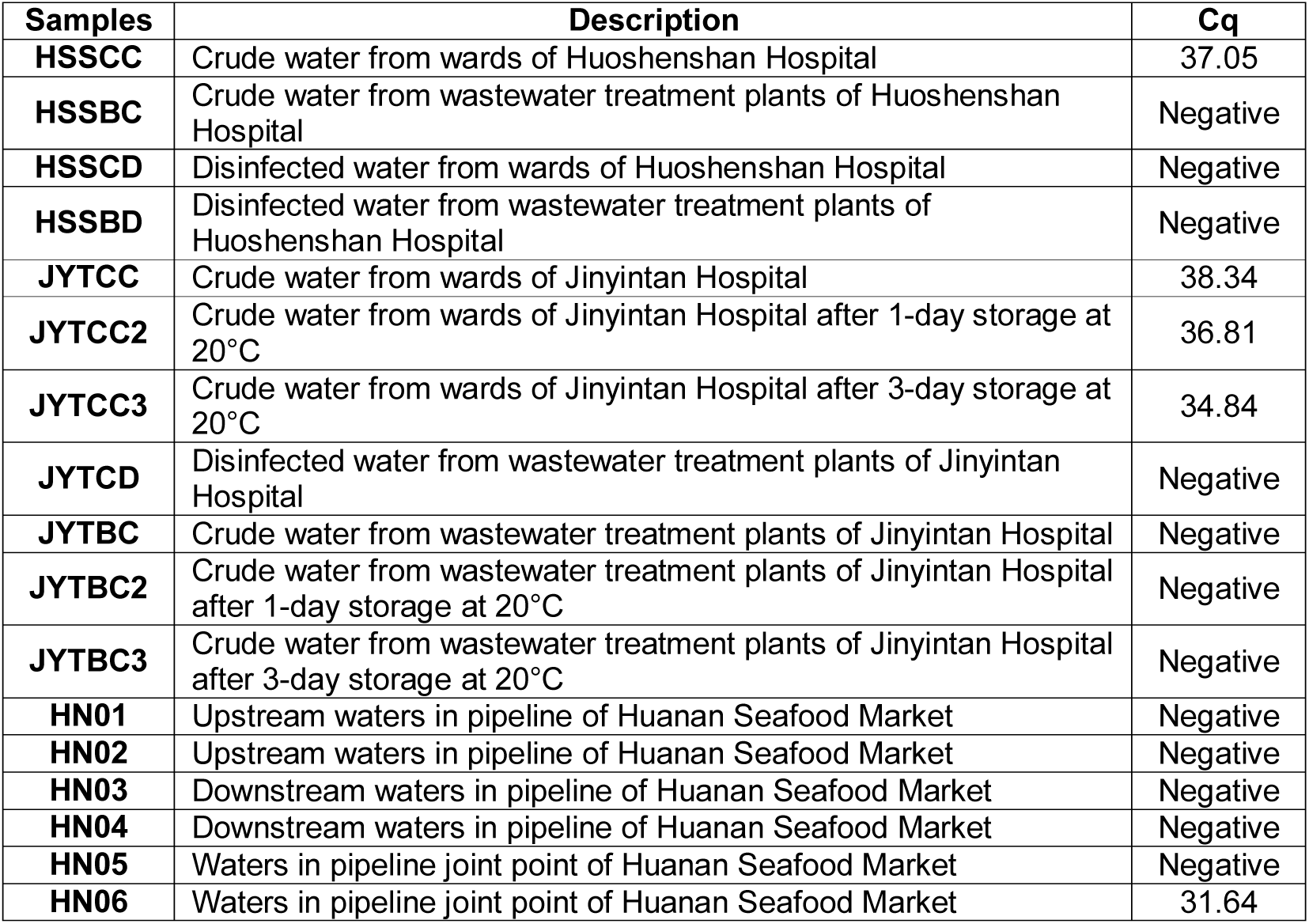
Sampling sites and quantification cycle (Cq) values of SARS-CoV-2 in water samples.

| Samples | Description | Cq |
| --- | --- | --- |
| HSSCC | Crude water from wards of Huoshenshan Hospital | 37.05 |
| HSSBC | Crude water from wastewater treatment plants of Huoshenshan Hospital | Negative |
| HSSCD | Disinfected water from wards of Huoshenshan Hospital | Negative |
| HSSBD | Disinfected water from wastewater treatment plants of Huoshenshan Hospital | Negative |
| JYTCC | Crude water from wards of Jinyintan Hospital | 38.34 |
| JYTCC2 | Crude water from wards of Jinyintan Hospital after 1-day storage at 20$^{\circ}$C | 36.81 |
| JYTCC3 | Crude water from wards of Jinyintan Hospital after 3-day storage at 20$^{\circ}$C | 34.84 |
| JYTCD | Disinfected water from wastewater treatment plants of Jinyintan Hospital | Negative |
| JYTBC | Crude water from wastewater treatment plants of Jinyintan Hospital | Negative |
| JYTBC2 | Crude water from wastewater treatment plants of Jinyintan Hospital after 1-day storage at 20$^{\circ}$C | Negative |
| JYTBC3 | Crude water from wastewater treatment plants of Jinyintan Hospital after 3-day storage at 20$^{\circ}$C | Negative |
| HN01 | Upstream waters in pipeline of Huanan Seafood Market | Negative |
| HN02 | Upstream waters in pipeline of Huanan Seafood Market | Negative |
| HN03 | Downstream waters in pipeline of Huanan Seafood Market | Negative |
| HN04 | Downstream waters in pipeline of Huanan Seafood Market | Negative |
| HN05 | Waters in pipeline joint point of Huanan Seafood Market | Negative |
| HN06 | Waters in pipeline joint point of Huanan Seafood Market | 31.64 |

### 2.2 Preparation of silver-nanorod SERS array

The aligned silver-nanorod SERS (SN-SERS) array was fabricated in oblique angle deposition (OAD) using a custom-designed electron beam/sputtering evaporation system (Suzhou Derivative Biotechnology Co., LTD.) and formed randomly on a 4-inch silicon wafer with increasing deposition time^35^. Briefly, Si-wafer was immersed in absolute alcohol and blow-dried up using N_2_ gas prior to loading on the substrate holder. The substrate holder was then fixed on the specially designed Glancing Angle Deposition (GLAD) sample stage in an e-beam evaporator. Deposition was performed at a base pressure lower than 3×10^−4^ Pa. The thickness of film growth was monitored using a quartz crystal microbalance. Firstly, a thin layer of about 20 nm was deposited to assist the adhesion of silver on Si-wafer, followed by the deposition of a base layer of 200 nm silver. The GLAD stage was then tilt to 84° with respect to the incident vapor. A layer of 80 nm was then deposited with substrate rotation at 0.1 rev/s to improve the seeding for nanorod growth. The deposition rate was 2 Å/s in each stage and lasted about 3 h.

### 2.3 Fabrication of ACE2@SN-SERS substrate

ACE2 was purchased from Novoprotein (China) and stored in borate buffer solution (0.1 M, pH=7.2) at −80°C before use. SN-SERS substrate was firstly cleaned by thorough rinse with deionized water and dried using N_2_ gas flow. Subsequently, 1 μL of ACE2 stock solution was loaded to SN-SERS substrate and placed in an incubator under constant temperature and humidity conditions (25°C; 75%, w/w) for 4 h. ACE2 was then bound onto the surface of SN-SERS substrate, designated as ACE2@SN-SERS substrate, which could be stored in 4°C for 2 weeks before use.

### 2.4 Raman spectral acquisition

For laboratory test, Raman spectra were acquired using a near-infrared confocal Raman microscope (HR evolution, Horiba, USA) equipped with a 785 nm near-IR laser source, a 300 l/mm grating and a semiconductor-cooling detector (CCD). All Raman spectra were collected with a 50× objective lens (NA=0.7) at an exposure time of 10 seconds, 3 accumulations, and laser power of 10 mW prior to lens. Raman spectroscopic system was calibrated with a silicon wafer at Raman shift of 520 cm^−1^. At least five random regions were measured for each sample, and a minimum of 9 individual spectra were acquired per sample. A 785-nm portable Raman spectrometer (Finder Edge, Zolix, China) was used for on-site diagnosis, and the ACE2@SN-SERS substrate was placed on the probe of the portable Raman spectrometer with the following parameters: 0.5 s acquisition time and 300 mW laser power. At least five-time measurement was conducted for each sample. For both laboratory and on-site test, all Raman spectra were recorded in the range of 100-3500 cm^−1^ in biological triplicates.

### 2.5 Multivariate analysis of Raman spectra

Raw spectral data were pre-processed by using the open source IRootLab toolbox performed on MATLAB r2012 ^36^ Briefly, each acquired Raman spectrum was cut to a biochemical-cell fingerprint region (900-1800 cm^−1^), baseline corrected, wavelet de-noised, and vector normalized. Unlike RT-qPCR and ELISA assay, our ACE2@SN-SERS assay generates Raman spectral data, which are multivariate and difficult to generate an individual variable for SARS-CoV-2. Thus, we used two approaches to distinguish the difference between positive and negative samples. Firstly, the ratio of Raman intensity at 1182 cm^−1^ to that at 1189 cm^−1^ was calculated, designated as 1182/1189 ratio, as an indicator for diagnostic prediction. Alternatively, multivariate analysis was applied to the pre-processed spectral data to reduce data dimensions and extract key information. Principal component analysis (PCA) is an unsupervised data analytical method reducing the dimensionality of data, determining principal components (PCs) and extracting key features ^37, 38, 39^. The first 10 PCs, which account for more than 90% of the variance of the selected spectral regions, were then inputted into linear discriminate analysis (LDA), which determines the discriminant function line that maximizes the inter-class distance and minimizes the intra-class distance to derive an optimal linear boundary separating the different classes^37^ Generally, PCA-LDA score plots and cluster vectors are generated, and the scores of the linear discriminant 1 (LD1) provides the best classification^39, 40^.

### 2.6 Statistical analysis

One-way ANOVA was used to compare the difference between samples and p-value less than 0.05 refers to statistically significant difference.

## 3. Results and discussions

### 3.1 Features of SN-SERS and ACE2@SN-SERS substrates

SEM images (Figure 2A) illustrated a successful fabrication of silver nanorods on SERS substrate. The overall diameter and length of silver nanorods was 211±45 and 737±52 nm, respectively, and the uniform structure demonstrated a density of 8 nanorods/μm^2^. After functionalization with ACE2 protein, clear protein structures were observed on ACE2@SN-SERS substrate, exhibiting as small islands with a diameter of 2 μm (Figure 2B).

**Figure 2.**
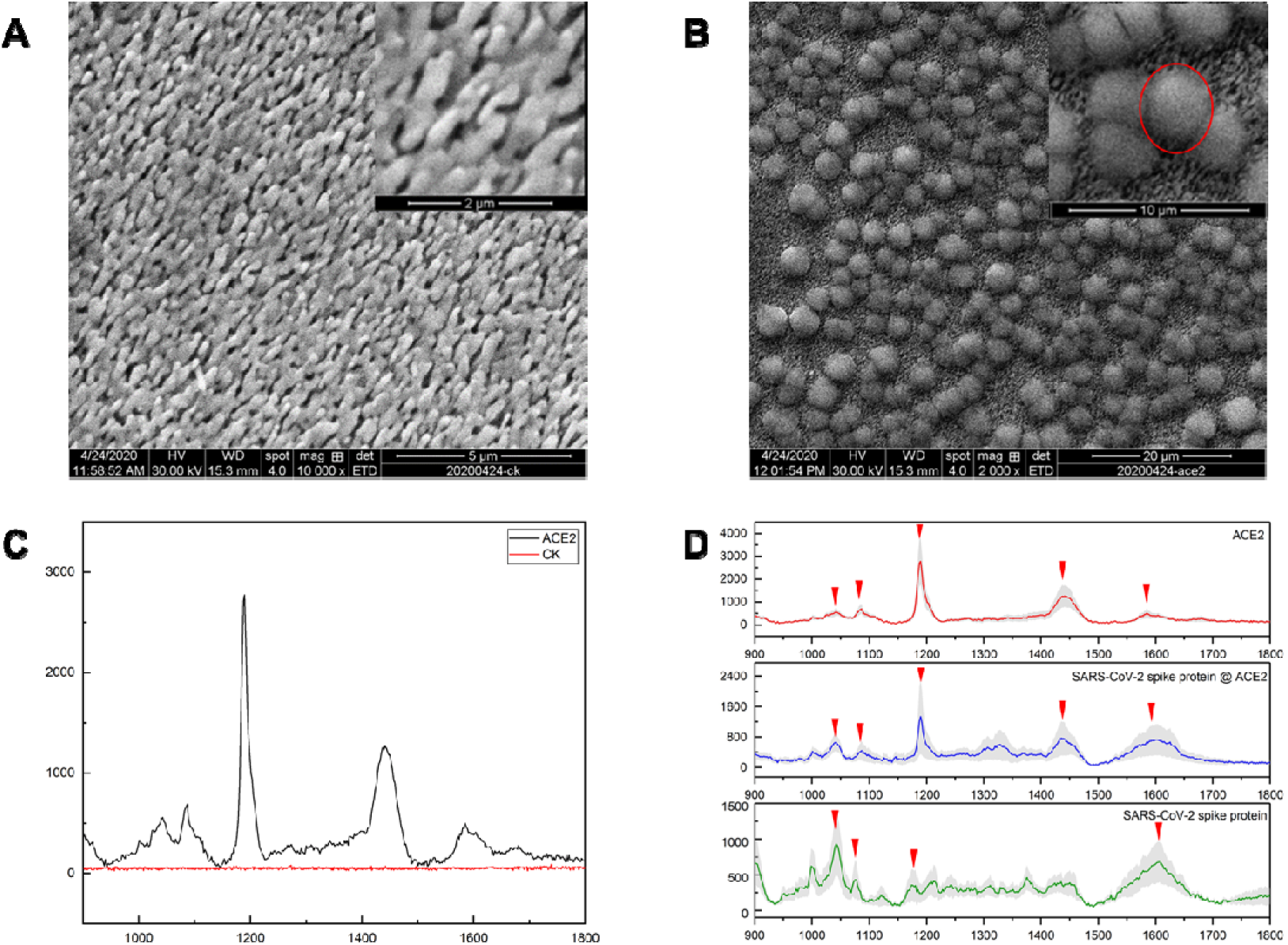
(A) SEM image of SN-SERS substrate. Scale bar: 5 μm (upper right: 2 μm). (B) SEM image of ACE2@SN-SERS substrate. Scale bar: 20 μm (upper right: 10 μm). Functionalized ACE2 protein is present as ‘islands’ (red circle). (C) SERS signals of SN-SERS substrate without ACE2-functionalization (CK) and ACE2@SN-SERS substrate (ACE2). (D) Comparison of SERS signals between ACE2@SN-SERS substrate, SARS-CoV-2 spike protein on SN-SERS substrate and SARS-CoV-2 spike protein on ACE2@SN-SERS substrate. Featured SERS signals of ACE2 protein include 1032, 1089, 1189, 1447 and 1587 cm^−1^.

The background Raman signals of SN-SERS substrate without ACE2 exhibited no significant peaks from 900 to 1800 cm^−1^ (Figure 2C), showing a satisfactory performance for interrogating bio-related SERS. ACE2@SN-SERS substrate generated remarkable SERS signals, around 400 times stronger than SN-SERS substrate (Figure 2C). The featured Raman peaks are probably assigned with phenylalanine (1032 cm^−1^)^41^, C-N stretching in protein (1089 cm^−1^)^42^, Amide III for C-N stretching and N-H bending (1189 cm^−1^)^43^, CH_2_ bending mode of proteins (1447 cm^−1^)^44^ and conjugated −C=C- in protein (1587 cm^−1^)^45^.

### 3.2 SERS detection of SARS-CoV-2 spike protein

SARS-CoV-2 spike protein was firstly tested on SN-SERS and ACE2@SN-SERS substrates as a proof-of-concept demonstration. Different from ACE2 protein, Raman peak intensities derived from SARS-CoV-2 spike protein were relatively weak on SN-SERS substrate and the distinct peaks were mostly located at 1032, 1051 and 1447 cm^−1^ (Figure 2D). They are probably assigned to phenylalanine (1032 cm^−1^)^41^, C-N stretching in protein (1051 cm^−1^)^42^, Amide III for C-N stretching and N-H bending (1189 cm^−1^)^43^, CH_2_ bending mode of proteins (1447 cm^-1^)^44^. After loading spike proteins onto ACE2@SN-SERS substrate, the whole spectra exhibited a quenching of SERS signal intensity, especially at Raman shifts of 1089, 1189 and 1447 cm^−1^ (Figure 2D). Particularly, a red-shift from 1189 to 1182 cm^−1^ representing N-H bending was observed. It is possibly induced by the change in N-H vibration mode or the bond length of N-H^46, 47^, in response to the interfered H-bond of ACE2 after recognizing SARS-CoV-2 spike protein. The crystal structure of SARS-CoV-2 spike RBD bound to the ACE2 receptor indicates the networks of hydrophilic interactions at RBD/ACE2 interfaces ^33^, which exhibits a conceivable existence of hydrogen bond elucidating the strength change of N-H bond ^38^ and explains the Raman spectral red-shift from1189 to 1182 cm^−1^. Thus, our results indicated that SARS-CoV-2 spike protein can be recognized and bound by ACE2@SN-SERS substrate, consequently changing ACE2 structure to induce the SERS signal quenching. Such significant Raman signal change demonstrated that our designed ACE2@SN-SERS assay has a satisfactory performance in SARS-CoV-2 interrogation.

### 3.3 Performance of ACE2@SN-SERS in interrogating SARS-CoV-2 in real water samples

In on-site test, Raman spectra of ACE2 acquired via the portable Raman spectrometer exhibited coherent spectral peaks presented at 1032, 1052, 1089, 1189, 1447 and 1527 cm^−1^ comparing to those from the research-level Raman spectrometer in laboratory (Figure 4A), although relatively high signal noises were observed. It indicated the robustness of our developed ACE2@SN-SERS array and feasibility for on-site diagnosis. Raman spectra of water samples on ACE2@SN-SERS array exhibited different SERS spectra (Figure 4A). Some of them illustrated significant signal quenching, *e.g*., JYTCC, JYTCC2 and JYTCC3, whereas others possessing similar spectral patterns as ACE2@SN-SERS array included HSSCD, HSSBD, etc.

RT-qPCR results had separated all water samples into positive and negative groups for SARS-CoV-2. To distinguish these two groups from SERS spectra, we first used 1189/1182 ratio as a biomarker, as the red-shift from 1189 to 1182 cm^−1^ was the most remarkable spectral alteration. Figure 3C shows a significant difference in 1189/1182 ratio between positive and negative groups (*p*<0.001), averagely 1.311±0.446 and 0.766±0.218, respectively. Setting 0.900 or 1.000 as the threshold, the accuracy of 1189/1182 ratio reaches 93.33%, and the false-positive and false-negative percentage is 10% and 0%, respectively (Figure 3E). Thus, 1189/1182 ratio is a satisfactory biomarker to diagnose the presence of SARS-CoV-2 in water samples.

**Figure 3.**
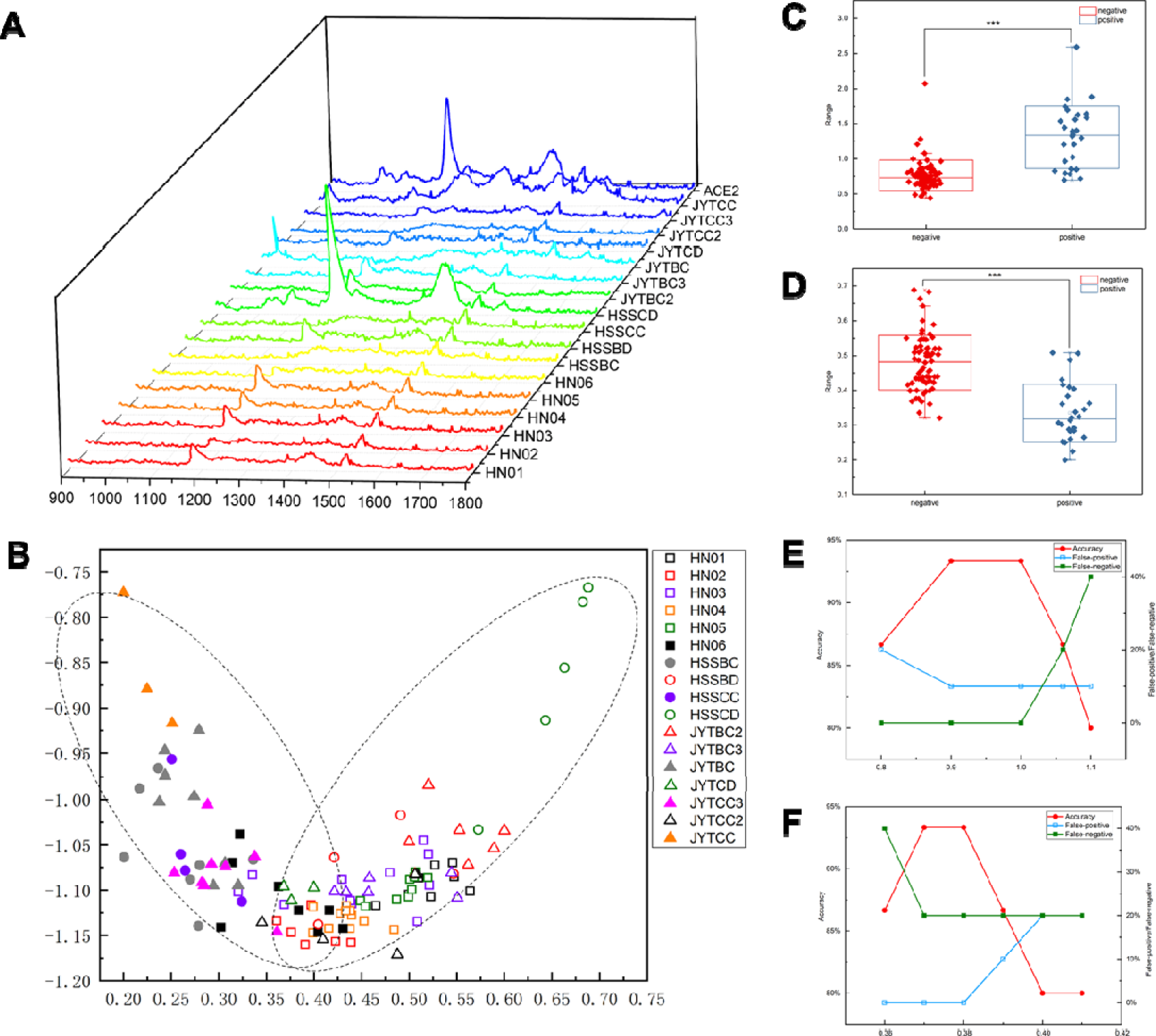
(A) SERS spectra of ACE2@SN-SERS and 17 tested water samples by ACE2@SN-SERS assay (mean value) using a portable Raman spectrometer. (B) Segregation of positive and negative water sample groups in PCA-LDA score plot. Solid and white dots represent positive and negative results for SARS-CoV-2, respectively. Grey dots refer to susceptive samples of HSSBC and JYTBC. (C) Difference of 1182/1189 ratio between positive and negative groups. (D) Difference of LD1 scores between positive and negative groups. (E) Accuracy, false positive and false-negative percentage based on 1182/1189 ratio. (F) Accuracy, false positive and false-negative percentage based on LD1 scores.

Nevertheless, the intrinsic SERS analysis provided convoluted Raman signals derived from ACE2 proteins or complexes of ACE2 and spike protein of SARS-CoV-2 located in plasmonic hot spot regions, it is crucial to employ multivariate method to extract information in these complex multivariable spectroscopic data for a better interrogation. Herein, the whole Raman spectra from 900 to 1800 cm^−1^ acquired Raman data from on-site test were analyzed by PCA-LDA in a hierarchical manner. PCA-LDA score plot clearly segregates the positive and negative groups regardless some overlaps (Figure 3B). The grey-labeled samples (HSSBC and JYTBC) are negative for SARS-CoV-2 by RT-PCR but fall into the positive group in PCA-LDA, whereas HN06 in PCA-LDA negative group had a Cq value of 31.61. Such contradictory results might be explained by different principles between these two assays. RT-qPCR targets SARS-CoV-2 RNA fragment, whereas our ACE2@SN-SERS assay recognizes SARS-CoV-2 spike protein. As RNA has a shorter half-life and is more easily degraded than protein, SARS-CoV-2 spike protein occur hypothetically longer time than RNA, particularly after disinfection could breakdown viral envelops and RNA. Since LD1 scores derived from PCA-LDA model provide the best classification, they are assigned as criteria and exhibit significant difference between the positive and negative groups (*p*<0.001). The accuracy by LD1 scores is 93.33% when the threshold is 0.370 or 0.380, while the false-positive or false-negative percentage is 0% and 20%, respectively. Together with 1182/1189 ratio, our findings proved the feasibility of the developed ACE2@SN-SERS assay to on-site interrogate SARS-CoV-2 in real water samples, and both indicators (1182/1189 ratio and LD1 scores) have satisfactory performances.

### 3.4 Presence of SARS-CoV-2 in real water samples from Wuhan

Cq value of crude water from wards of Huoshenshan Hospital (HSSCC) and Jinyintan Hospital (JYTCC) was 37.05 and 38.34, respectively, showing the presence of SARS-CoV-2 viral RNA. The results became negative in the biological treatment sector (HSSBC) and aerobic biodegradation sector (JYTBC) in the wastewater treatment plants of Huoshenshan and Jinyintan Hospitals (Table 1). However, both 1182/1189 ratios and LD1 scores by ACE2@SN-SERS assay had contradictory results that SARS-CoV-2 was present in all wastewater samples throughout in the treatment process despite of slight decay (Figure 4A and 4E). It might be explained by higher stability of SARS-CoV-2 spike protein than RNA after disinfection, and the residual spike proteins were still detectable but the infectivity was of low risk. Our findings indicated a decay of SARS-CoV-2 viral RNA along wastewater treatment process, consistent with previous reported facts that both RNA (Norovirus GGI, GGII, sapovirus, and Aichi virus) ^48, 49, 50^ and DNA viruses (enteric adenoviruses, JC polyomaviruses, BK polyomaviruses) ^50^ were effectively removed in conventional and biological wastewater treatment plants. The presence of SARS-CoV-2 in adjusting tanks might pose threats to workers for medical wastewater treatment, and their personal care protections are suggested to prevent potential infection. The rapidness and easy operation of this developed ACE2@SN-SERS assay offers a solution to monitor SARS-CoV-2 in medical wastewater treatment plants, allowing on-site assessment of potential spreading risks on workers and surrounding environment.

**Figure 4.**
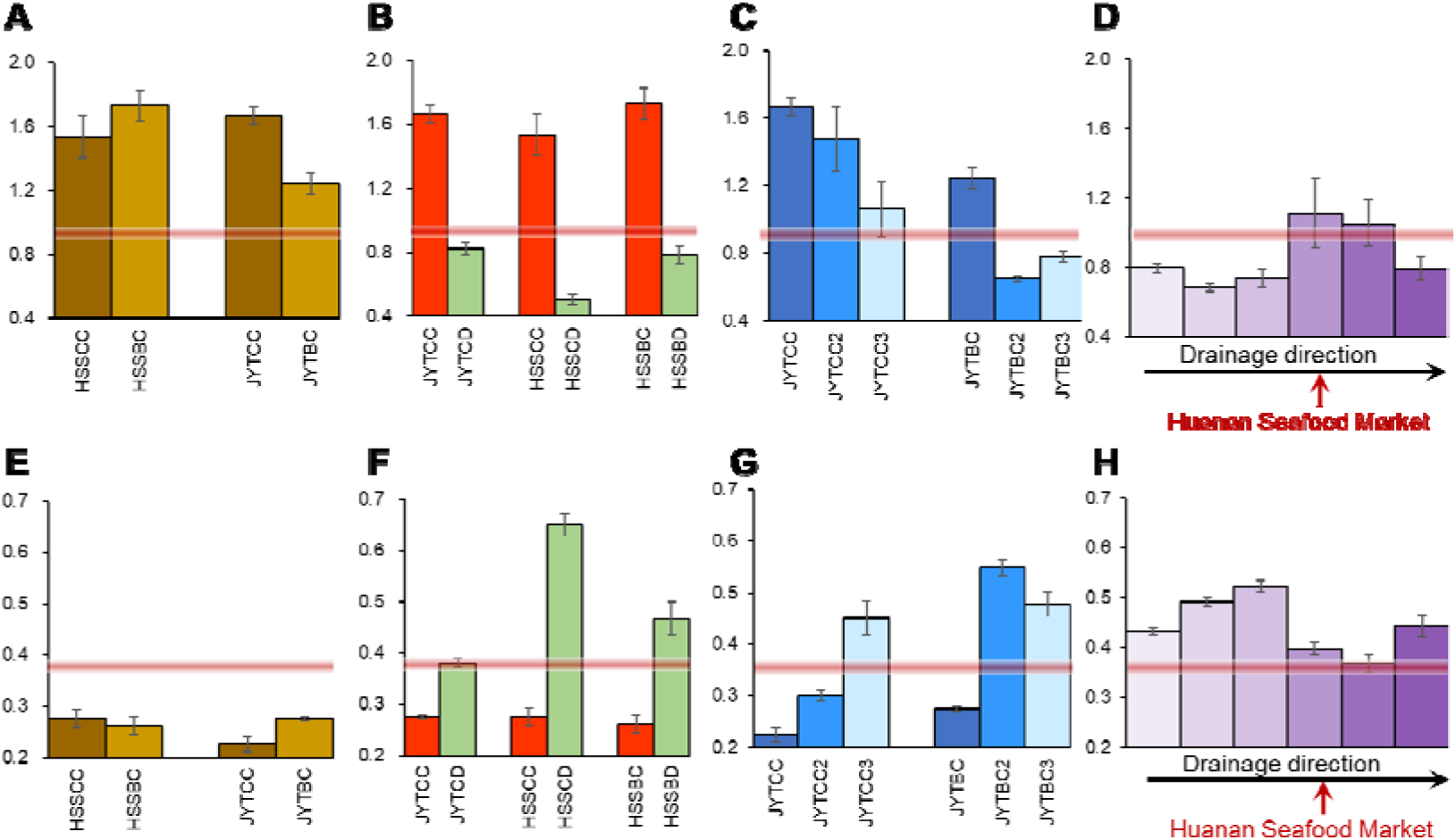
Applications of on-site SERS interrogation for SARS-CoV-2. Change of 1189/1182 ratio (A) and LD1 score (E) along wastewater treatment process for wastewater treatment plant management. Significant difference of 1189/1182 ratio (B) and LD1 score (F) between crude and disinfected waters for determination of disinfection efficiency. Change of 1189/1182 ratio (C) and LD1 score (G) for viral survival in environmental media. Occurrence of SARS-CoV-2 in the pipeline of Huanan Seafood Market by 1189/1182 ratio (D) and LD1 score (H).

Disinfection with 500 mg/L of sodium hypochlorite completely removed SARS-CoV-2 viral RNA, exhibiting negative RT-qPCR and ACE2@SN-SERS results for disinfected water from wards of Huoshenshan Hospital (HSSCD), wastewater treatment plant of Huoshenshan Hospital (HSSBD), and wards of Jinyintan Hospital (JYTCD) (Figure 4B and 4F). These results documented the robustness of ACE2@SN-SERS assay and matched well with a previous study that coronaviruses was efficiently inactivated by surface disinfection procedures ^51^.

SARS-CoV-2 was positive in JYTCC2 (1-day storage at 20°C) and JYTCC3 (3-day storage at 20°C) as evidenced by both RT-qPCR and ACE2@SN-SERS assays (Figure 4C), suggesting SARS-CoV-2 viral RNA or spike protein could persist in medical wastewater for at least 3 day (Figure 4C and 4G). SARS-CoV-2 could survive longer on plastic and stainless steel than copper and cardboard, up to 72 hours ^52^. Our results provided evidence using two assays to show the prolonged presence of SARS-CoV-2 in medical wastewater.

Diagnosing patients and tracking asymptomatic candidates is one of major challenges for COVID-19 prevention and control. In this work, we collected water samples from pipeline receiving discharge from Huanan Seafood Market, a suspected first place of COVID-19 outbreak in China. The upstream waters in pipeline around Huanan Seafood Market demonstrated negative RT-qPCR results for SARS-CoV-2 viral RNA, and exhibited no SARS-CoV-2 spike protein from clear Raman shifts at 1189 cm^−1^ in ACE2@SN-SERS assay (Figure 3A). After receiving wastewater from Huanan Seafood Market, the positive RT-qPCR results (Table 1, Cq=31.64), 1182/1189 ratios (Figure 4D) and LD1 scores (Figure 4H) hinted the entry of SARS-CoV-2 into pipelines, and it declined in downstream pipe network. Different from RT-qPCR assay requiring RNA extraction and laboratory amplification, the developed ACE2@SN-SERS assay allows onsite diagnosis of SARS-CoV-2 with portable Raman spectrometers, showing huge potentials for field practices to track the presence and source of SARS-CoV-2 in sewage pipe network.

### 3.5 Prospective

In this study, our developed ACE2@SN-SERS assay provides good accuracy and satisfactory performance for the rapid and onsite diagnosis of SARS-CoV-2 in environmental specimens, meeting well with RT-qPCR results. Compared with RT-qPCR and ELISA assays, this ACE2@SN-SERS assay has three advantages. Firstly, ACE2@SN-SERS assay does not rely on RNA extraction or immune biomarker, simplifying the sample preparation procedure and shortening the measurement time. Secondly, spike protein is more stable than RNA as the biomarker for SARS-CoV-2, hinting a more stable and sensitive assay particularly for environmental specimens. Last but not the least, we have proved that portable Raman spectrometers can provide satisfactory signals with ACE2@SN-SERS assay and allows rapid interrogation of SARS-CoV-2 on site.

There are some limitations in this work. Firstly, our ACE2@SN-SERS assay cannot evaluate viral viability or infectivity as free spike proteins or viral envelop are also recognizable by ACE2@SN-SERS substrates to quench SERS signals, possibly overestimating the presence of SARS-CoV-2 in human or environmental specimens. Secondly, only limited real samples were tested and the interference of other viruses targeting ACE2 for cell entry is still questionable, owing to the restriction in sample collection and clinical tests during the outbreak of COVID-19 in Wuhan. Nevertheless, the mentioned advantages prove ACE2@SN-SERS assay as a reliable and for mobile detection platform or screening system for clinal and environmental diagnosis onsite under a variety of conditions. Our state-of-the-art work also raises a concept to screen other viruses with RBD recognizing receptors of human cells for entry and evaluate the recognition strength of SARS-like viruses across mammalian species, when different RBD-containing viruses are tested or human cell receptors are functionalized for substrate fabrication. As a possible in-vitro assay, ACE2@SN-SERS substrate might also contribute to the assessment of vaccine efficiency. Further studies need to address those possibilities and establish robust databases and algorithms for a faster interrogation, even down to 1 min, for clinical and environmental purposes.

## 4. Conclusion

A novel ACE2@SN-SERS assay was developed in this study by functionalizing human cellular receptor ACE2 proteins on silver-nanorods and generating strong SERS signals. The successful and significant quenching of SERS signal intensities in the presence of SARS-CoV-2 spike proteins proved its capability in capturing and recognizing SARS-CoV-2. Onsite tests on 17 water samples using a portable Raman spectrometer achieved satisfactory performance in interrogating the presence of SARS-CoV-2 in environmental specimens, although some inconsistent with RT-qPCR results. At the current stage, the developed ACE2@SN-SERS assay had acceptable accuracy, false-positive and false-negative percentages, which can be further improved by fabrication implementation, database set-up and algorithm optimization. It has a bright future and huge potential as a rapid and on-site diagnostic tool for SARS-CoV-2 and other viruses to confirm patients, determine community cases and track environmental viral sources in pandemics.

## Data Availability

All data are available by request.

## 5. Acknowledgement

The authors would like to thank the project from Ministry of Science and Technology of the People’s Republic of China (2020YFC0842500) for providing water samples, and Science and Technology Service Network Initiatives (KFJ-STS-QYZX-061) and Scientific Research Equipment Development Project (YZ201653) for SN-SERS substrate fabrication.

## 6. Author contributions

Concept and design: DZ.

Laboratory data acquisition: XZ, RM, YZ, JL.

On-site data acquisition: DZ, XZ.

Analysis or interpretation of data: DZ, SD, XW, XH, YL, GL, JQ.

Drafting of the manuscript: DZ, JL.

Statistical analysis: DZ, JL.

## Notes

### Competing Interest Statement

The authors have declared no competing interest.

